## Supplemental Document for "Individualized Machine-learning-based Clinical Assessment Recommendation System"

### 1. Dataset

#### *1.1. Synthetic Dataset Generation*

The basis for creating synthetic datasets 1-3 is the scenario highlighted in Supplementary Figure 1. We created a condition where two additional features display equally good predictive power as analyzed by SHAP. However, a global feature selection is shown to be insufficient as it only selects one out of the two equally good features to add for all scenarios. This limitation can lead to suboptimal inference since it overlooks the information provided by the initial feature. Synthetic datasets 1-3 are thus designed to illustrate the necessity of incorporating individualized feature recommendations, demonstrating that considering only one feature for all is inadequate for accurate inference.

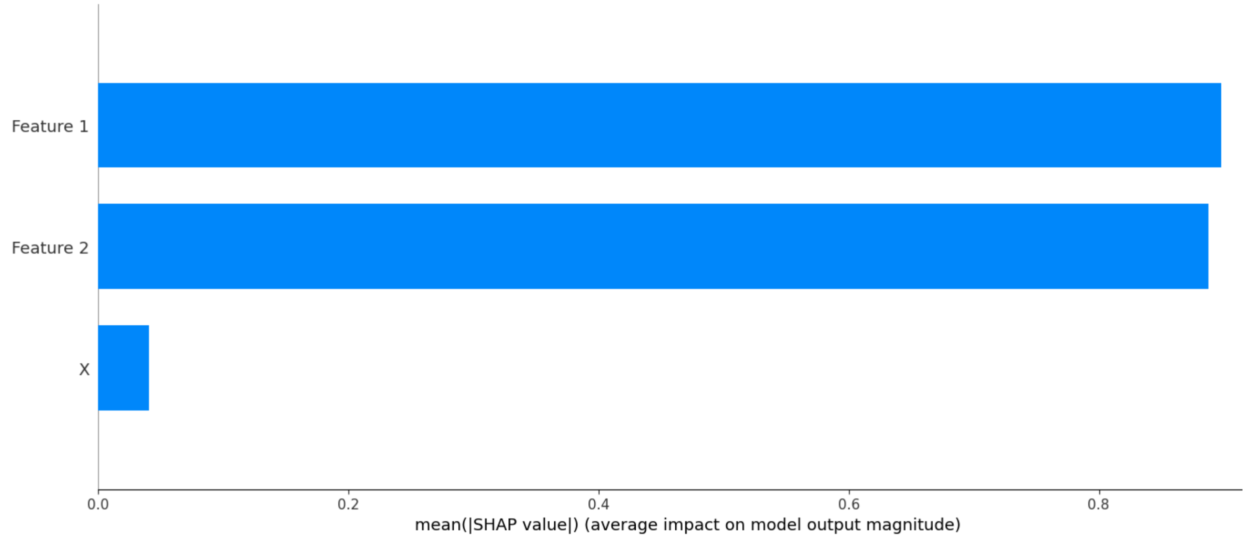


Figure 1: **SHAP values bar graph for Feature 1 and Feature 2.** The X-axis represents the SHAP value (feature importance measure) for each feature on the Y-axis. X on the Y-axis represents the initial feature set that the patient started with. The graph highlights the comparable

magnitudes for SHAP values associated with 'Feature 1' and 'Feature 2,' emphasizing the challenge faced by global feature selection methods. Traditional approaches will fall short and pick either Feature 1 or Feature 2 to add to X without any rigorous selection criteria (i.e., random selection). However, a personalized machine learning framework can adapt to individual observation and add features according to the information in X.

We created synthetic dataset 1 to have two equally good features. However, the predictiveness of each feature operates on different patient groups. Feature 1 is set to be a good predictor for X below 0.5, and Feature 2 is set to be a good predictor for X above 0.5. The variable X is the initial feature value.

---

**Synthetic Dataset 1**

---

```
Initialize X, y, feature_1, feature_2
X ← random values ranging from 0 to 1
y ← random values either 0 or 1
Generate Feature 1:
for all x_i, y_i ∈ X, y
    if x_i > 0.5 then feature_1[i] ← random values ranging from 0 to 1
    else
        if y_i = 0 then feature_1[i] ← random values ranging from 0 to 0.5
        if y_i = 1 then feature_1[i] ← random values ranging from 0.5 to 1
Generate Feature 2:
for all x_i, y_i ∈ X, y
    if x_i < 0.5 then feature_2[i] ← random values ranging from 0 to 1
    else
        if y_i = 0 then feature_2[i] ← random values ranging from 0 to 0.5
        if y_i = 1 then feature_2[i] ← random values ranging from 0.5 to 1
```

For synthetic dataset 2, Feature 1 is set to be a good predictor for X below 0.5, and Feature 2 is set to be a good predictor for X above 0.5, similar to synthetic dataset 1. However, the decision space is non-linear. For example, for X < 0.25, Feature 1 can be used to decide the outcome where Feature 1 < 0.5 means outcome = 0 and Feature 2 > 0.5 means outcome = 1. However, this decision boundary is flipped for 0.25 < X < 0.5 where Feature 1 < 0.5 means outcome = 1 and Feature 2 > 0.5 means outcome = 0.

---

**Synthetic Dataset 2**

---

```
Initialize X, y, feature_1, feature_2
X ← random values ranging from 0 to 1
```

```
    y ← random values either 0 or 1
    Generate Feature 1:
    for all x_i, y_i ∈ X, y
        if x_i > 0.5 then feature_1[i] ← random values ranging from 0 to 1
        else if x_i < 0.25
            if y_i = 0 then feature_1[i] ← random values ranging from 0 to 0.5
            if y_i = 1 then feature_1[i] ← random values ranging from 0.5 to 1
        else if x_i < 0.5
            if y_i = 1 then feature_1[i] ← random values ranging from 0 to 0.5
            if y_i = 0 then feature_1[i] ← random values ranging from 0.5 to 1
    Generate Feature 2:
    for all x_i, y_i ∈ X, y
        if x_i < 0.5 then feature_2[i] ← random values ranging from 0 to 1
        else if x_i > 0.75
            if y_i = 0 then feature_2[i] ← random values ranging from 0 to 0.5
            if y_i = 1 then feature_2[i] ← random values ranging from 0.5 to 1
        else if x_i > 0.5
            if y_i = 1 then feature_2[i] ← random values ranging from 0 to 0.5
            if y_i = 0 then feature_2[i] ← random values ranging from 0.5 to 1
```

For synthetic dataset 3, Feature 1 is set to be a good predictor for X below 0.7, and Feature 2 is set to be a good predictor for X above 0.3. This creates an overlapping predictive region where any additional feature is a good addition.

---

**Synthetic Dataset 3**

---

```
    Initialize X, y, feature_1, feature_2
    X ← random values ranging from 0 to 1
    y ← random values either 0 or 1
    Generate Feature 1:
    for all x_i, y_i ∈ X, y
        if x_i > 0.7 then feature_1[i] ← random values ranging from 0 to 1
        else
            if y_i = 0 then feature_1[i] ← random values ranging from 0 to 0.5
            if y_i = 1 then feature_1[i] ← random values ranging from 0.5 to 1
    Generate Feature 2:
    for all x_i, y_i ∈ X, y
        if x_i < 0.3 then feature_2[i] ← random values ranging from 0 to 1
        else
            if y_i = 0 then feature_2[i] ← random values ranging from 0 to 0.5
```

**if** *y_i* = 1 **then** *feature_2[i]* ← random values ranging from 0.5 to 1

For synthetic dataset 4, Feature 1 is set to be a good predictor for all X values, and Feature 2 is set to be a good predictor for all X values. This creates a condition where personalization is unnecessary, as either feature can be globally selected and still produce a good prediction.

---

**Synthetic Dataset 4**

---

**Initialize** *X, y, feature_1, feature_2*
*X* ← random values ranging from 0 to 1
*y* ← random values either 0 or 1
**Generate Feature 1:**
**for all** *x_i, y_i* ∈ *X, y*
  **if** *y_i* = 0 **then** *feature_1[i]* ← random values ranging from 0 to 0.5
  **if** *y_i* = 1 **then** *feature_1[i]* ← random values ranging from 0.5 to 1
**Generate Feature 2:**
**for all** *x_i, y_i* ∈ *X, y*
  **if** *y_i* = 0 **then** *feature_2[i]* ← random values ranging from 0 to 0.5
  **if** *y_i* = 1 **then** *feature_2[i]* ← random values ranging from 0.5 to 1

For synthetic dataset 5, Feature 1 is set to be a good predictor for all X values while Feature 2 and 3 are set to be bad predictors for all X values. This creates a condition where personalization is not necessary, as a global feature selection will pinpoint Feature 1's usefulness and use it for all samples.

---

**Synthetic Dataset 5**

---

**Initialize** *X, y, feature_1, feature_2, feature_3*
*X* ← random values ranging from 0 to 1
*y* ← random values either 0 or 1
**Generate Feature 1:**
**for all** *x_i, y_i* ∈ *X, y*
  **if** *y_i* = 0 **then** *feature_1[i]* ← random values ranging from 0 to 0.5
  **if** *y_i* = 1 **then** *feature_1[i]* ← random values ranging from 0.5 to 1
**Generate Feature 2:**
**for all** *x_i, y_i* ∈ *X, y*
  *feature_2[i]* ← random values ranging from 0 to 1
**Generate Feature 3:**
**for all** *x_i, y_i* ∈ *X, y*
  *feature_3[i]* ← random values ranging from 0 to 1

*1.2. Real-World Dataset Preparation*

To ensure relevance and manage computational complexity, datasets were selected based on the following criteria: binary classification problem type, containing 10 to 100 features, and comprising 100 to 1000 samples. The early diabetes dataset comprises 16 features and 520 samples, while the heart failure dataset comprises 11 features and 299 samples. For the early diabetes dataset, all of the features are binary values except age. Therefore, we applied normalization to the ‘age’ feature so that its values range from 0 to 1. For the heart failure dataset, we normalize all features as they are continuous. We removed some entries with missing values. We also removed the ‘time’ column as it was unnecessary for inference.

---

**Early Diabetes Preprocessing**

---

**Initialize** *df*<br>
*df* ← Read csv file 'diabetes_data_upload.csv'<br>
**Normalize Age Feature:**<br>
**for all** *age* ∈ *df[‘Age’]*<br>
    **do** min-max normalization **on** *age*<br>
**update** *age* values in *df*<br>
**save** *df* to 'early_diabetes_normalized.csv'

---

**Heart Failure Preprocessing**

---

**Initialize** *df*<br>
*df* ← fetch file from ‘ucirepo(id=519)’<br>
**Normalize All Feature:**<br>
**for all** *feature* ∈ *df*<br>
    **do** min-max normalization **on** *feature*<br>
**update** values in *df*<br>
**Remove Missing Values:**<br>
**for all** *entries* ∈ *df*<br>
    **if** *entries* have missing values **then**<br>
        **delete** *entries*<br>
**Remove ‘Time’ Column:**<br>
**delete** *df[‘time’]*<br>
**save** *df* to 'heartfailure_normalized.csv'

### 2. Code for Running iCARE Experiment

**Algorithm 1:** *train_base_model(dataframe, target, key)*

```
Initialize X, y
X ← dataframe without target column
y ← dataframe[target]
Split Dataset into Train and Test Set:
X_train, y_train ← random 80% split of X, y using key as random seed
X_test, y_test ← the rest 20% of X, y
Train Logistic Regression Model:
lr ← LogisticRegression model fitted with X_train and y_train
Calculate Accuracy and AUC:
y_pred ← Generate prediction using lr on X_test
y_prob ← Generate probability using lr on X_test
acc ← calculate accuracy score using y_pred and y_test
auc ← calculate auc score using y_prob and y_test
return acc, auc
```

**Algorithm 2:** *train_lw_model(dataframe, target, key)*

```
Initialize X, y
X ← dataframe without target column
y ← dataframe[target]
Split Dataset into Train and Test Set:
X_train, y_train ← random 80% split of X, y using key as random seed
X_test, y_test ← the rest 20% of X, y
Calculate Sample Weights for Test Set:
for all x_i ∈ X_test
        distances[i] ← Euclidean distances between x_i and all sample in X_train
for all distance_i ∈ distances
        weights[i] ← if (distances_i != 0) then (1/distances_i) else 1
Train a Model for Each Test Sample:
for all weights_i ∈ weights
        lr[i] ← LogisticRegression model fitted with X_train and y_train with weights_i
Calculate Accuracy and AUC:
for all x_i, lr_i ∈ X_test, lr
y_pred[i] ← Generate prediction using lr_i on X_i
```

```
y_prob[i] ← Generate probability using lr_i on X_i
acc ← calculate accuracy score using y_pred and y_test
auc ← calculate auc score using y_prob and y_test
return acc, auc
```

**Algorithm 3:** *train_global_model(dataframe, initial_features, target, key)*

```
Initialize X, y
X ← dataframe without target column
y ← dataframe[target]
Split Dataset into Train and Test Set:
X_train, y_train ← random 80% split of X, y using key as random seed
X_test, y_test ← the rest 20% of X, y
Train Logistic Regression Model:
lr ← LogisticRegression model fitted with X_train and y_train
Find Most Important Feature:
explainer ← SHAPLinearExplainer using lr on X_train
shap_values ← generate SHAP values using explainer on X_train
for all shap_i ∈ shap_values
    shap_values[i] ← mean absolute of SHAP values in shap_i
best_feature ← feature corresponding to index i where shap_values[i] is maximum
Create a Dataset with only a Subset of Features:
if best_feature not in initial_feature
    add best_feature to initial_features
    dataframe_with_recommendation ← dataframe with initial_features columns
else
    dataframe_with_recommendation ← dataframe with initial_features columns
Train a Model Based on the Dataset with Recommendation:
acc, auc ← train_base_model(dataframe_with_recommendation, target, key)
acc_lw, auc_lw ← train_lw_model(dataframe_with_recommendation, target, key)
return acc, auc, acc_lw, auc_lw
```

**Algorithm 4:** *train_pml(dataframe, initial_features, target, key)*

```
Initialize X, y
X ← dataframe without target column
y ← dataframe[target]
Split Dataset into Train and Test Set:
```

```
    X_train, y_train ← random 80% split of X, y using key as random seed
    X_test, y_test ← random 20% split of X, y using key as random seed
    Create a Dataset with only a Subset of Features:
    X_train_subset, y_train_subset ← random 80% split of X, y using key as random seed,
                                        only initial_features columns present
    X_test_subset, y_test_subset ← the rest 20% of X, y,
                                        only initial_features columns present
    Calculate Sample Weights for Test Set:
    for all x_i ∈ X_test_subset
        distances[i] ← Euclidean distances between x_i and all sample in X_train_subset
    for all distance_i ∈ distances
        weights[i] ← if (distances_i != 0) then (1/distances_i) else 1
    Train a Model with All Features for Each Test Sample:
    for all weights_i ∈ weights
        lr[i] ← LogisticRegression model fitted with X_train and y_train with weights_i
    Find Most Important Feature for Each Model:
    best_features = []
    for all lr_i ∈ lr
        explainer ← SHAPLinearExplainer using lr on X_train
        shap_values ← generate SHAP values using explainer on X_train
        for all shap_i ∈ shap_values
        shap_values[i] ← mean absolute of SHAP values in shap_i
        best_feature[i] ← feature corresponding to index i where
                                   shap_values[i] is maximum
    Train Using Recommendation:
    for all recommendation_i ∈ best_features
        test_sample ← X_test[i] with appended recommendation_i feature value
        new_dataset ← X_train_subset, y_train_subset appended with recommendation_i
                                   feature value
        lr_i ← LogisticRegression model fitted with new_dataset
        lr_i_lw ← LogisticRegression model fitted with new_dataset with weights_i
        y_pred[i] ← Generate prediction using lr_i on test_sample
        y_prob[i] ← Generate probability using lr_i on test_sample
        y_pred_lw[i] ← Generate prediction using lr_i_lw on test_sample
        y_prob_lw[i] ← Generate probability using lr_i_lw on test_sample
    Calculate Accuracy and AUC:
    acc, auc ← calculate accuracy and auc score using y_pred, y_prob and y_test
    acc_lw, auc_lw ← calculate accuracy and auc score using y_pred_lw, y_prob_lw and
                      y_test
    return acc, auc, acc_lw, auc_lw
```

**Algorithm 5:** *run_experiment(dataframe, initial_features, target, key)*

**Iterate 100 times:**
**for** *key* **in** *range(100)*
    **Generate Ceiling Model Performance:**
    *ceiling_acc[key], ceiling_auc[key]* ← *train_base_model(dataframe, target, 0)*
    **Generate Global Model Performance:**
    *global_acc[key], global_auc[key], global_acc_lw[key], global_auc_lw[key],* ←
        *train_global_model(dataframe, initial_features, target, key)*
    **Generate iCARE Model Performance:**
    *pml_acc[key], pml_auc[key], pml_acc_lw[key], pml_auc_lw[key],* ←
        *train_global_model(dataframe, initial_features, target, key)*
**Calculate Mean:**
*ceiling_acc_mean* ← mean of *ceiling_acc*
*ceiling_auc_mean* ← mean of *ceiling_auc*
*global_acc_mean* ← mean of *global_acc*
*global_auc_mean* ← mean of *global_auc*
*global_acc_lw_mean* ← mean of *global_acc_lw*
*global_auc_lw_mean* ← mean of *global_auc_lw*
*pml_acc_mean* ← mean of *pml_acc*
*pml_auc_mean* ← mean of *pml_auc*
*pml_acc_lw_mean* ← mean of *pml_acc_lw*
*pml_auc_lw_mean* ← mean of *pml_auc_lw*
**return** *ceiling_acc_mean, ceiling_auc_mean, global_acc_mean, global_auc_mean,*
    *global_acc_lw_mean, global_auc_lw_mean, pml_acc_mean, pml_auc_mean,*
    *pml_acc_lw_mean, pml_auc_lw_mean*

### 3. Code Availability

The implementation of the iCARE framework can be found in the following github link: https://github.com/DevinRS/iCARE. Code is written in Python 3.11.9.
